# Variant-specific SARS-CoV-2 within-host kinetics

**DOI:** 10.1101/2021.05.26.21257835

**Authors:** Baptiste Elie, Bénédicte Roquebert, Mircea T. Sofonea, Sabine Trombert-Paolantoni, Vincent Foulongne, Jérémie Guedj, Stpéhanie Haim-Boukobza, Samuel Alizon

## Abstract

Since early 2021, SARS-CoV-2 variants of concern (VOCs) have been causing epidemic rebounds in many countries. Their properties are well characterised at the epidemiological level but the potential underlying within-host determinants remain poorly understood. We analyse a longitudinal cohort of 6,944 individuals with 14,304 cycle threshold (Ct) values of qPCR VOC screening tests performed in the general population and hospitals in France between February 6 and August 21, 2021. To convert Ct values into numbers of virus copies, we performed an additional analysis using droplet digital PCR (ddPCR). We find that the number of viral genome copies reaches a higher peak value and has a slower decay rate in infections caused by Alpha variant compared to that caused by historical lineages. Following the evidence that viral genome copies in upper respiratory tract swabs are informative on contagiousness, we show that the kinetics of the Alpha variant translate into significantly higher transmission potentials, especially in older populations. Finally, comparing infections caused by the Alpha and Delta variants, we find no significant difference in the peak viral copy number. These results highlight that some of the differences between variants may be detected in virus load variations.

## Introduction

SARS-CoV-2 ‘Variants of Concern’ (VOC) correspond to lineage that cause phenotypically different infections from ‘historical’ lineages with increases in contagiousness [1, 2, 3, 4, 5] and virulence [1, 6, 7]. Because of the deadly epidemic rebounds they caused, they are closely monitored through full genome sequencing but also targeted Reverse-Transcription Polymerase Chain Reaction (RT-PCR) screening. The latter is less precise than the former but more affordable, allowing for wider testing [8]. Those assays yield a quantitative value, the cycle threshold (Ct), which is often used as a proxy for the amount of virus genetic material [9, 10], although this metric should be handled with care for coronaviruses [11].

Many studies analysed the epidemiology of VOCs but their within-host properties are less clear. Indeed, cross-sectional studies alone are not sufficient to characterise potential variants impacts on virus dynamics due to identifiability issues [12, 13, 14]. Still, a few cross-sectional studies have reported lower Ct values for Alpha than for ‘historical’ infections [1, 15] suggesting that VOCs could be causing infections with higher virus loads. Therefore, longitudinal analyses are necessary to better understand the within-host kinetics of SARS-CoV2 variants infections.

The field of within-host kinetics has grown focusing mainly on chronic infections, but some studies consider acute infections [16]. In the case of SARS-CoV-2, several studies used Ct values as a proxy for virus load to report temporal variations within individuals, on longitudinal data from hospitalized patients [17], health workers [18], and experimental infections in non-human primates [19]. More recently, studies focusing on the effect of the vaccine showed an overall decrease in viral loads among vaccinated infected patients [20], or a faster viral load decrease [21, 22], although this remains unclear for symptomatic infections [23]. These studies illustrate that viral kinetics reflect some properties of the infection and can explain some of the variations in the detection probability.

Here, we analyse SARS-CoV-2 kinetics in a large number of individuals using linear mixed models, which allows us to explore how virus load dynamics may vary depending on the setting (general population or hospitals) or the variant causing the infection.

## Material and Methods

### Data

This study was approved by the CHU of Montpellier’s Institutional Review Board and is registered at ClinicalTrials.gov (n° NCT04653844). The data originates from variant-specific RT-PCR tests performed in France between February 06 and August 21, 2021 on SARS-CoV-2 positive samples [15, 13].

The multiplex assays use probes targeting variant-specific mutations (see Table 1), as well as a region in the N virus gene for control purposes. We use the Ct of the later probe in our analyses.

**Table 1:** Summary of the assays used to screen for VOCs among positive tests. WT stands for ‘wild type’, i.e. ancestral lineages.

| Assay | Detailed name | Targets | Variants |
| --- | --- | --- | --- |
| IDS1 | ID<sup>TM</sup> SARS-CoV-2/UK/SA Variant Triplex (ID Solution) | N501Y, Δ69-70 | WT/$\delta$ vs. $\alpha$ vs. $\beta/\gamma$ |
| IDS2 | ID<sup>TM</sup> SARS-CoV-2 / N501Y/ E484K Quadruplex (ID Solution) | N501Y, E484K | WT/$\delta$ vs. $\alpha$ vs. $\beta/\gamma$ |
| IDS3 | ID<sup>TM</sup> SARS-CoV-2 / VOC evolution Pentaplex (ID Solution) | L452R, E484K, E484Q | WT/$\alpha$ vs. $\beta/\gamma$ vs. $\delta$ |
| Perkin | VariantDetect<sup>TM</sup> SARS-CoV-2 RT-PCR (PerkinElmer) | L452R, E484K, E484Q | WT/$\alpha$ vs. $\beta/\gamma$ vs. $\delta$ |

Given the specificity of each assay used, different formatting steps were used. In particular, the initial IDS1 test designed to distinguish between the historical lineages and the Alpha VOC may increase the proportion of the latter VOC for high Ct values. Further details are shown in the flow diagram in Figure 1 and can be found in earlier studies [8, 15, 13].

**Figure 1:**
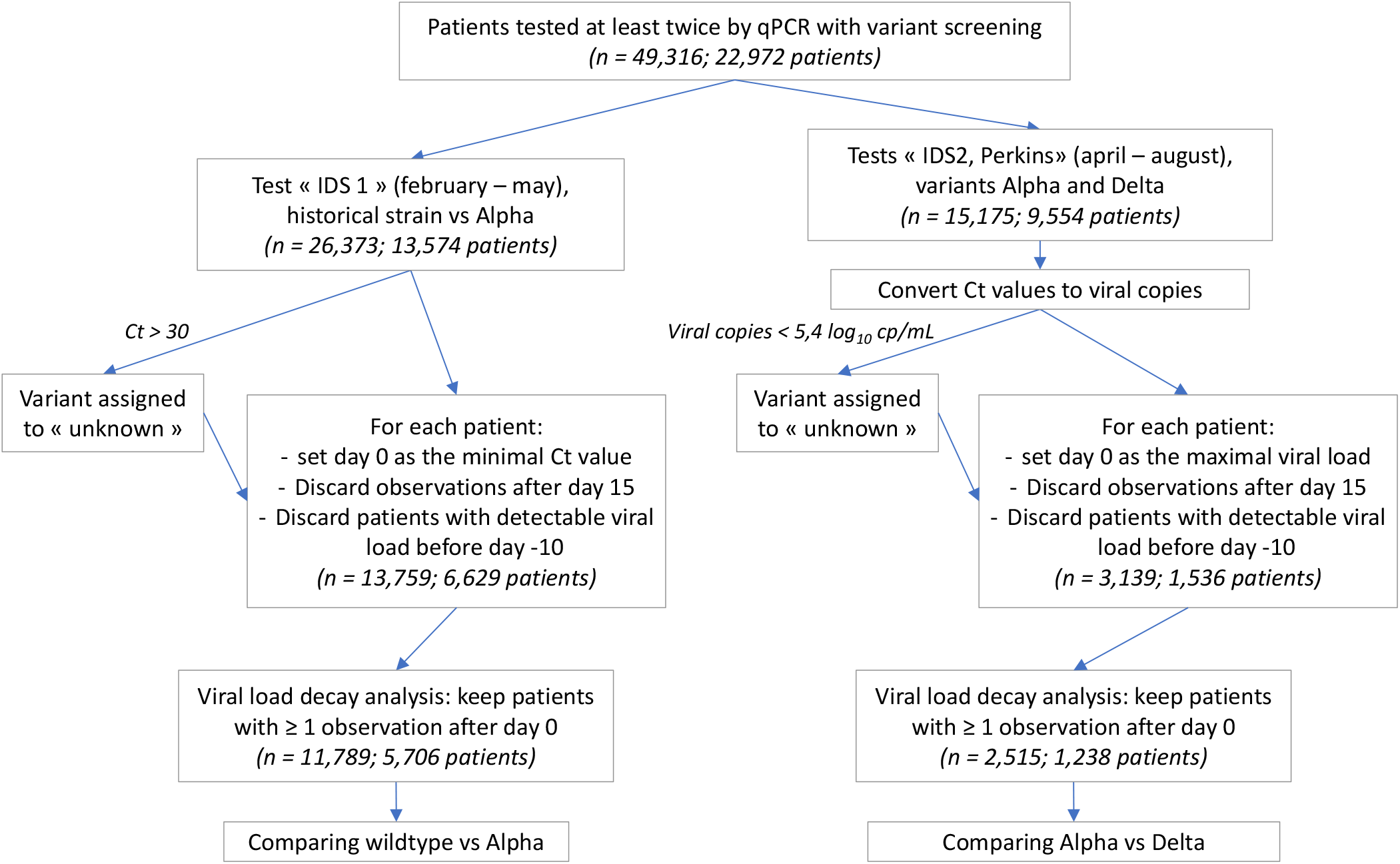
Dataset formatting steps. *n* indicates the number of tests, i.e. Ct values, analysed. For the IDS1 assay, the analyses were performed directly on the Ct values because the provider did not have any remaining tests to use with the ddPCR calibration. Furthermore, the assessment of the virus lineage (i.e. wild type or VOC) was only performed for tests with Ct values lower or equal than 30. For results obtained using the IDS2 and Perkin assays, Ct values were converted into viral genome copies before subsequent analysis and the assessment of the virus lineage was only performed for tests with more than 5.4*log*_10_ copies/mL.

### Digital droplet PCR

Using Ct measures as a proxy for virus load has several limitations, especially in the case of Coronaviruses [11]. Here, we do not attempt to equate the two but rather assume that temporal variations in Ct values are associated in changes in infectiousness.

To further investigate the biological significance of Ct values, we analysed samples from infections by a known virus lineage, i.e. historical or VOC, using both a variant-specific PCR and a digital droplet PCR (ddPCR). More precisely, we used two couples of primers targeting regions N1 and N2 in the virus, and the cellular human RNAse P for internal control.

### Statistical analyses

#### Linear mixed models

We used a linear model to study the potential link between the Ct value of the screening test and the number of virus copies obtained using ddPCR, using virus lineage and test assay as a co-factor. The results of the statistical model were used to convert Ct values from all assays into a number of virus copies.

We analysed the longitudinal data of number of virus copies (or Ct values for IDS1) with linear mixed models and used the R package lme4 [24] to fit the restricted maximum likelihood parameters to the data. The response variable was the number of virus copies (or the Ct value for IDS1), and we included two random effects on the individual and on the region of sampling.

To select which additional effects to include in the linear mixed model, we compared models with all possible parameters combinations listed in Table 2. The best model was chosen based on the Akaike Information Criterion (AIC) [25].

**Table 2:** Effects included in the models being tested.

| Effect | Values | Details |
| --- | --- | --- |
| Virus strain | Historical, Alpha, Delta | Variant screening test result |
| Day | 0-15 days | Day 0 being that with the lowest Ct value in an individual |
| Hospitalization | Yes or No | If the patient was sampled at least once in an hospital. |
| Age | 5-97 | Age of the patient |
| Variant reproduction number | 0.7-1.8 | Average number of secondary infections caused by an infected person at a given date [26], stratified by region. |
| Delay between 1<sup>st</sup> and 2<sup>nd</sup> test | 1-15 days | Proxy for the bias in the time of first test, assuming 2<sup>nd</sup> test is done 7 days after symptoms onset. |
| Vaccine coverage | 0-100% | Vaccine coverage at the time of infection for the corresponding age class. |
| Interaction between day and each effect |  | This represents the impact of each effect on the viral copy number decay after peak. |
| Interaction between the age and virus strain |  | This represents the differential impact of the variant on the viral copy number peak in function of the age. |

We verified that models with Δ_*AIC*_ *<* 2 were qualitatively identical to the model with the best AIC [25]. We also verified that the censored data points (*i*.*e*. the Ct values above the limit of detection, which was set at 37, or the viral copies number below 10,000 copies/mL) did not have a significant impact on the results by computing all the linear mixed models selected by a Δ_*AIC*_ *<* 2 with a censored effect using the lmec R package.

Differences in viral copies between populations from the linear model outputs were statistically assessed using the Tukey adjustment and the emmeans R package.

#### Variant specific reproduction number

We first calculated the global epidemic reproduction number in each French region using hospital admission data [27]. We then adjusted this number by using the estimated relative proportion of each variant among the daily infections caused [5, 8, 15], and the mean transmission advantage computed by an independent study [28].

#### Transmission potential

We used the infectiousness profile estimated from the data of [29] corrected by [30], *i*.*e*. a shifted Gamma distribution, with shape 97.19, rate 3.72, and shift 25.63.

This analysis being restricted to one test (IDS1), we studied the correlation between the Ct value and the instantaneous infectiousness, *i*.*e*. the infectiousness profile density.

We then used this mapping to infer a transmission potential, which can be seen as a proxy for the basic reproduction number [31]. Using the outputs of our model capturing within-patient dynamics, we integrated the infectiousness obtained from the Ct values from day 0 to day 15.

## Data availability

Data and code are available on https://gitlab.in2p3.fr/ete/sars-cov2_kinetics

## Results

### From Ct values to number of virus copies

For a given RT-PCR assay, we found a log-linear relationship between the Ct value and the absolute number of viral copies measured by ddPCR (Figure 2). Compared to the reference (*i*.*e*. IDS2), IDS3 yielded a Ct value 1.96 lower (Student t-test, Tukey adjustment: *p* < 10^−4^), and Perkin a Ct value 6.9 higher (*p* < 10^−4^). For the Perkin assay, we also detected a significant effect of the variant on the Ct value. For the same number of virus copies, we found a Δ*Ct* of 5.0 between samples originating from an infection by a Delta or an Alpha variant (*p* < 10^−4^).

**Figure 2:**
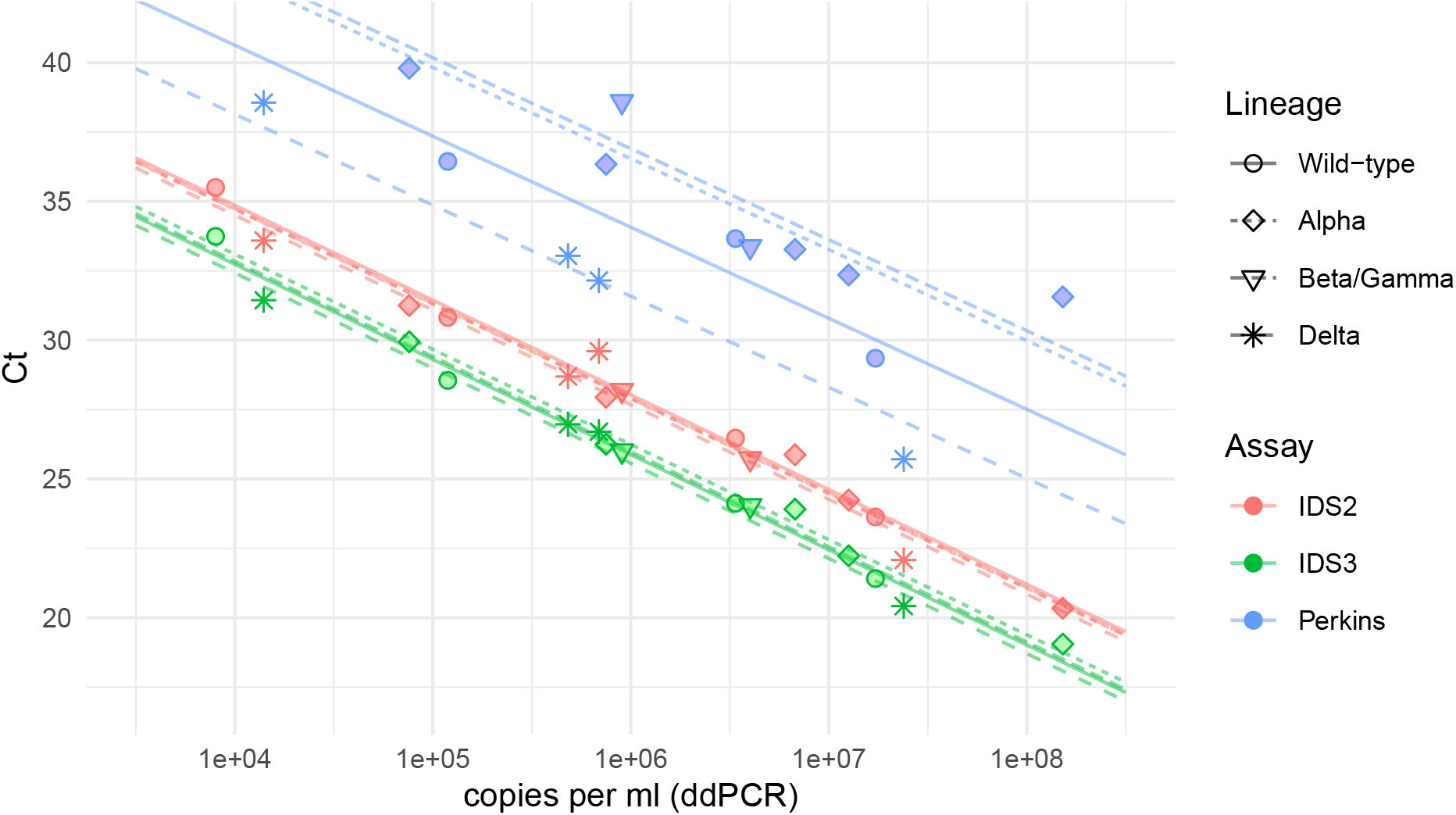
Ct value as a function of SARS-CoV2 copy number. The shape indicates the virus lineage and the color the variant screening assay used to obtain the Ct value for the control probe targeting the N gene. Virus copy number was estimated using a digital droplet PCR targeting the N gene.

### Historical lineages vs. the Alpha variant

After formatting the database for the IDS1 assay (Figure 1), we identified 12,536 suitable samples (Table 3). Due to the limited number of observations, we removed patients infected lineages consistent with the Beta/Eta/Gamma VOCs.

**Table 3:** Properties of the longitudinal datasets: i) based on the ISD1 assay (Historical lineages vs. Alpha variant), the data contain 12,536 Ct values from 6,064 individuals, and ii) based on IDS2, Perkin or IDS3 assays (Alpha vs. Delta variants), the data contain 2,751 Ct values from 1,239 individuals.

| Variable | Value | Historical strains vs. Alpha variant model<br>Median (IQR) or n % | Alpah vs. Delta variant model<br>Median (IQR) or n % |
| --- | --- | --- | --- |
| sampling context | *general population (ref)* | 5706 (94%) | 1263 (93%) |
|  | *hospital* | 358 (6%) | 92 (7%) |
| age |  | 41 [25-58] y.o. | 35 [23-54] y.o. |
| follow up duration |  | 8 [6-10] days | 6 [5-7] days |
| date of first sampling (in 2021) |  | Mar 9 [Feb 20 -Mar 22] | May 5 [Apr21 -Jul21] |
| lineage | *wildtype* | 1211 (20%) (*ref*) | 6 (0.4%) |
|  | *Alpha* | 4495 (74%) | 849 (63%) (*ref*) |
|  | *Beta/Gamma/Eta* | 358 (6%) | 110 (8%) |
|  | *Delta* | 0 | 390 (29%) |
| samples per individual | 2 | 5686 (93%) | 1313 (97%) |
|  | >2 | 378 (7%) | 42 (3%) |
| administrative region | *Ile-de-France* | 3863 (64%) | 676 (50%) |
|  | *Normandie* | 659 (11%) | 181 (13%) |
|  | *Hauts-de-France* | 612 (10%) | 142 (10%) |
|  | *PACA* | 273 (4%) | 204 (15%) |
|  | *other* | 657 (11%) | 151 (12%) |
| Screening assay | *IDS1* | 12,536 | 0 |
|  | *IDS2* | 0 | 1,928 (70%) |
|  | *Perkin* | 0 | 824 (30%) |
|  | *IDS3* | 0 | 1 (<1%) |

To analyze the virus load kinetics, we inferred linear mixed models and selected the one with the lowest AIC. The estimated values of the model are shown in Table 4.

**Table 4:** Linear mixed model parameters estimates. Bold rows correspond to estimates with p < 0.05. The notation *a*:*b* indicates an interaction between factors *a* and *b*. CI stands for ‘confidence interval’. The random effect of the patients on the intercept has a standard deviation of 2.075 [1.95,2.19], the random effect of the region on the intercept has a standard deviation of 1.051 [0.61,1.72] and the residues have a standard deviation of 3.89 [3.82,3.96].

| predictor |  | Estimate | 95% CI |
| --- | --- | --- | --- |
| **intercept** |  | **24.9** | **(24.1,25.7)** |
| **day** |  | **1.25** | **(1.20,1.30)** |
| age |  | -0.0049 | (-0.011,0.001) |
| hospitalisation | *no* | ref | — |
|  | *yes* | **-1.36** | **(-1.75,-0.97)** |
| delay between first and second test |  | **-0.40** | **(-0.43,-0.37)** |
| **day:age** |  | **-0.0029** | **(-0.0036,-0.0022)** |
| day:strain | *Historical* | ref | — |
|  | *Alpha* | **-0.059** | **(-0.097,-0.021)** |
| age:strain | *Historical* | ref | — |
|  | *Alpha* | **-0.016** | **(-0.020,-0.011)** |

The linear mixed model revealed significant differences in Ct values dynamics between the historical lineages and the Alpha VOC (Figure 3 A). First, compared to historical lineages infections, the peak Ct appeared to decrease with age for infections caused by the Alpha VOC (Table 4). Overall, for the French age structure, the peak Ct was significantly lower (−0.67 Ct [-0.87,-0.46]). Furthermore, in infections caused by the Alpha VOC, the rate of Ct increase over the infection was lower. As a result, 7 days after the viral copy number peak, the Ct difference was larger (−1.08 Ct [-1.32,-0.85]). Finally, we also observed a significant impact of the hospitalization status, with a lower peak Ct value.

**Figure 3:**
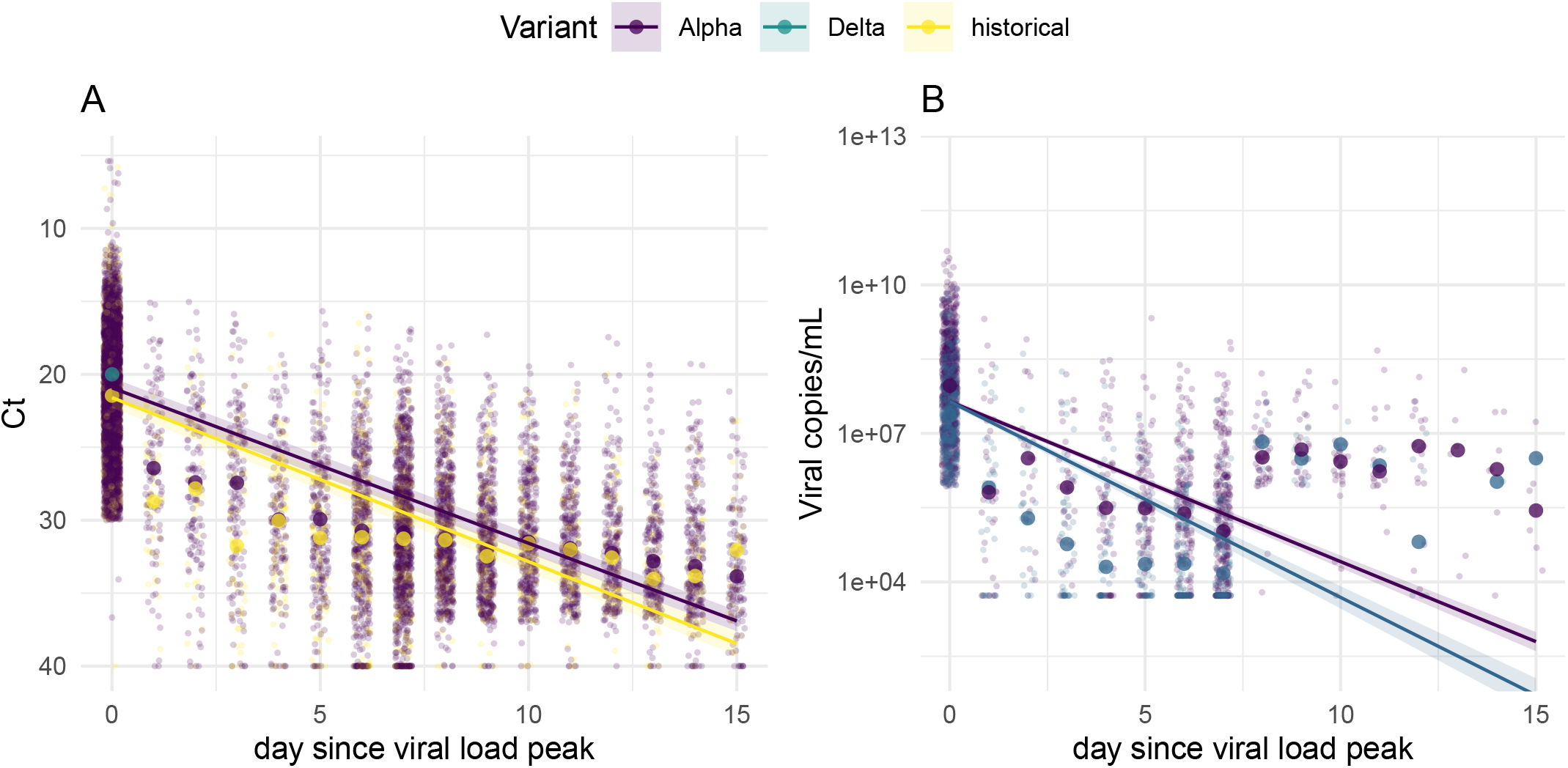
Within-host longitudinal Ct data as a function of the virus lineage. The dots represent the observed values. The bold dots represent the median value for each day and each strain. The lines represent the linear model for an average patient (median age, non hospitalised). A. Model with Ct values of historical lineages vs. the Alpha variant. B. Model of virus copies per mL of Alpha vs. Delta variant. Both model were set approximately on the same scale on the y-axis.

### Alpha transmission potential advantage

To further investigate the implications of these results at the population level, we performed a mapping between the daily infectivity and the daily Ct value after its peak for the wild type strain inferred from the linear model. The resulting significant correlation (Figure 4A, *R*^2^ = 0.95) supports a linear relationship between Ct and infectiousness.

**Figure 4:**
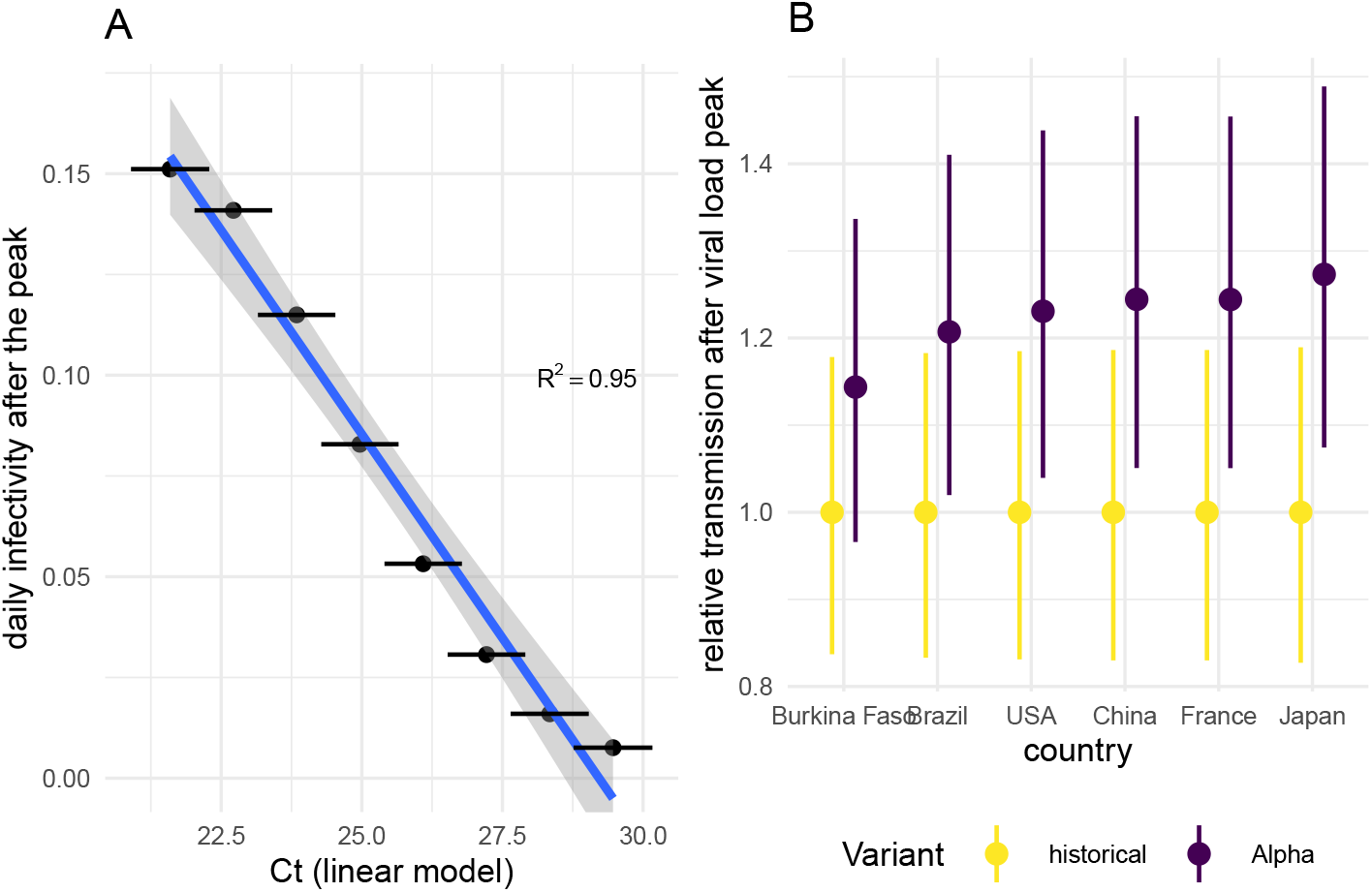
Impact of the differential viral genome copies on the transmissibility. A. Relationship between infectivity and Ct, after the viral genome copies peak, for the historical strain. B. The linear models parameters were used, with the demography of each country, to infer the transmission advantage of the variants. The error bars represent the 95% bootstrap quantile.

We used this mapping between Ct values and infectivity to study transmission potential differences between lineages. We found that the advantage of the Alpha VOC over the historical strain was more pronounced in countries with older populations (Figure 4B).

### Alpha vs. Delta variant

We then analysed the data which involved Ct values from 3 other assays that we were able to convert into virus copy numbers (see the Methods and Figure 2). In the following, we use a log base 2 relationship with time since peak of viral genome copies to compare the orders of magnitude with the previous results.

Infections consistent with viruses from Beta/Gamma/Eta lineages were too rare to be analysed and removed them from the dataset. Overall, we were able to compare infections caused by the Alpha or the Delta variant by analyzing the remaining 2,515 samples (Table 3). Compared to the previous dataset, patients were slightly younger (median age 35 vs 41) and the follow-up duration was shorter (median 6 vs 8 days). This latter difference was corrected by the variable taking into account the delay between first and second test.

The best model according to the AIC criterion is detailed in Table 5 and the dynamics of the number of viral genome copies is shown in Figure 3 B. On 15 out of 17 models with a Δ*AIC <* 2, we found no significant difference between the kinetics of the infections caused by the Alpha and the Delta VOC on the peak viral copy number. However, we found a significant difference on the decay rate on 13 models.

**Table 5:** Linear model parameters estimates. Bold rows correspond to estimates with p<0.05. CI stands for ‘confidence interval’. The notation *a*:*b* indicates an interaction between factors *a* and *b*. The random effect of the patients on the intercept has a standard deviation of 1.69 [1.42, 1.91], and the residues have a standard deviation of 3.50 [3.37,3.64].

| Predictor |  | Estimate | 95% CI |
| --- | --- | --- | --- |
| **Intercept** |  | **20.14** | **(18.9,21.4)** |
| **day** |  | **-1.33** | **(-1.42,-1.24)** |
|  | age | **0.024** | **(0.013,0.034)** |
| R(t) variant specific |  | 0.90 | (-0.19,1.98) |
| **Vaccine coverage** |  | **-0.034** | **(-0.045,-0.023)** |
| **Delay between first and second test** |  | **0.67** | **(0.59,0.74)** |
| **day:age** |  | **0.0061** | **( 0.0043,0.0080)** |
| **day:hospital** |  | **0.12** | **(0.0018,0.25)** |
| day:strain | *Alpha* | ref | — |
|  | *Delta* | **-0.24** | **(-0.33,-0.15)** |

To make sure that this result was not due to confounding factors, we included several covariates in our analysis which revealed a strong effect of the sampling date, which could interfere with the variant effect (since the Delta VOC rapidly replaced the Alpha VOC). The vaccine coverage in the population, which was used a proxy for the probability that the individual was vaccinated, as well as the delay between first and second test, measuring behavioural differences, were retained as a cofactor in the best linear model. Furthermore, the variant-specific reproduction number *R*(*t*), estimated at the regional level in France was also retained as a significant covariate, which is consistent with previous observations [13, 14], although the effect was not statistically significant.

## Discussion

Understanding the within-host kinetics of SARS-CoV-2 infections yielded original insights on infection virulence [17], or efficiency of screening strategies [18]. We analyzed a large national dataset of longitudinal RT-PCR Ct values to test the hypothesis that epidemic rebounds associated with SARS-CoV-2 variants could be linked to specificites in their within-host kinetics.

A linear mixed model indicated that infections caused by SARS-CoV-2 Alpha variant have higher virus loads, with a significant age-dependence. Furthermore, the temporal decrease in virus load was found to be slower when infections were caused by Alpha instead of the historical lineages. The results are consistent with results from a different cohort in France [32].

To further investigate the consequences of these variations in within-host kinetics at the epidemiological level, we assumed a linear correlation between Ct value and daily infectiousness, which is consistent with an earlier study [33] and confirmed by a modelling approach [34]. Translating the estimated Ct kinetics into transmission potential profiles revealed that the high viral copy number observed in Alpha VOC infections were consistent with a 25% increase in transmission potential compared to historical strains.

Kinetics of samples collected in hospitals exhibited higher peak viral genome copies (i.e. lower Ct values), which is consistent with earlier studies [17]. The results were unaffected by the removal of hospital data from the analysis.

We were unable to convert the Ct values from the first assays into number of virus genomes, which precluded us from comparing the Delta variant to the ancestral lineages. Therefore, we limited our comparison of the Delta variant to the infections caused by the Alpha variant. Our models show no difference in the peak of viral copy number, but they detect a faster decrease in infections caused by the Delta variant than in that caused by the Alpha variant. This may appear as counter-intuitive given the latter has a clear transmission advantage over the former [5, 28]. This could be explained by the fact that ddPCR itself is limited because it does not count the number of infectious virions [11]. Indeed, experiments suggest that for a given Ct value, samples from Delta VOC infections have a higher number of infectious particle than that from Alpha VOC infections [35]. Finally, this result is also consistent with others that find little differences in Ct values between infections caused by the Alpha and the Delta VOC [22].

A limitation of this analysis is that we do not have any indication regarding the date of the infection or of the symptom onset. This uncertainty prevented us from analyzing more mechanistic models with nonlinear mixed-effect models [17]. However, since the nature of the virus causing the infection is unlikely to affect the number of days between infection and screening, we do not expect our assumption that the lowest Ct value corresponds to the peak viral genome copies to introduce biases.

Our comparison between the Alpha and Delta VOCs is potentially subject to additional biases. This part of the analysis covers a large period of sampling, with an important increase in the vaccine coverage, and potential behavioural changes. We attempted to correct for those biases by including covariates such as vaccine coverage in the general population, variant reproduction number, or the delay between the first two tests of an infected individual. In particular, the use of antigenic tests as a first line of screening is likely to have delayed the delay between infection and qPCR testing. Their use increased from 20% to 50% of the total Covid-19 screening tests between May and August in France [36]. We observe that the median delay between the first and the second test (which is usually done 7 days after the symptoms) decreased from 7 to 5 days during the analysis. Therefore, we interpret a shorter delay as a first test done later on the infection, yielding lower viral copies number.

Overall, our study illustrates the insights that the combination of large screening data and statistical analyses can bring to the understanding of within-patient kinetics and population spread, as illustrated by our comparison between the Alpha variant and ancestral lineages. It also shows the limitations of Ct values and the added value of infectivity assays.

## Data Availability

Data will be available online on a gitlab repository

https://gitlab.in2p3.fr/ete/sars-cov2_kinetics

## Acknowledgements

The authors acknowledge the ISO 9001 certified IRD i-Trop HPC (South Green Platform) at IRD montpellier for providing HPC resources that have contributed to the research results reported within this paper.

We thank all the ETE modeling team for thoughtful discussions.

## Authors contribution

MTS, SHB, BR, and SA conceived the project, STP, VF, SHB, and BR collected the data, BE, MTS, and SA analysed the data, JG provided statistical expertise, BE and SA wrote the first draft the manuscript, and all authors contributed to the final version of the manuscript.

